# “Monozygotic twins discordant for severe clinical recurrence of COVID-19 show drastically distinct T cell responses to SARS-Cov-2”

**DOI:** 10.1101/2021.03.26.21253645

**Authors:** Mateus V. de Castro, Keity S. Santos, Juliana S. Apostolico, Edgar R. Fernandes, Rafael R. Almeida, Gabriel Levin, Jhosiene Y. Magawa, João Paulo S. Nunes, Miriam Bruni, Marcio M. Yamamoto, Ariane C. Lima, Monize V. R. Silva, Larissa R. B. Matos, Vivian R. Coria, Erick C. Castelli, Marilia O. Scliar, Andreia Kuramoto, Fernanda R. Bruno, Lucas C. Jacintho, Kelly Nunes, Jaqueline Y. T. Wang, Veronica P. Coelho, Miguel Mitne Neto, Rui M. B. Maciel, Michel S. Naslavsky, Maria Rita Passos-Bueno, Silvia B. Boscardin, Daniela S. Rosa, Jorge Kalil, Mayana Zatz, Edecio Cunha-Neto

## Abstract

**Background:** Clinical recurrence of COVID-19 in convalescent patients has been reported, which immune mechanisms have not been thoroughly investigated. Presence of neutralizing antibodies suggests other types of immune response are involved.

**Methods:** We assessed the innate type I/III IFN response, T cell responses to SARS-CoV-2 with IFNγ ELISPOT, binding and neutralizing antibody assays, in two monozygotic twin pairs with one COVID-19 recurrence case.

**Results:** In pair 1, four months after a first mild episode of infection for both siblings, one displayed severe clinical recurrence of COVID-19. Twin pair 2 of siblings underwent non-recurring asymptomatic infection. All fours individuals presented similar overall responses, except for remarkably difference found in specific cellular responses. Recurring sibling presented a reduced number of recognized T cell epitopes as compared to the other three including her non-recurring sibling.

**Conclusions:** Our results suggest that an effective SARS-CoV-2-specific T cell immune response is key for complete viral control and avoidance of clinical recurrence of COVID-19. Besides, adaptive immunity can be distinct in MZ twins. Given the rising concern about SARS-CoV-2 variants that evade neutralizing antibodies elicited by vaccination or infection, our study stresses the importance of T cell responses in protection against recurrence/reinfection.

**Key points:** Immune parameters leading to COVID-19 recurrence/reinfection are incompletely understood. A COVID-19 recurrence case in a monozygotic twin pair is described with an intact antibody and innate type I/III Interferon response and drastically reduced number of recognized SARS-CoV-2 T cell epitopes.

## BACKGROUND

Coronavirus SARS-Cov2, the causative agent of COVID-19 severe acute respiratory syndrome, has caused the most critical public health crisis in the last 100 years (1). Clinical recurrence of PCR-confirmed COVID-19 in persons with previous COVID-19 infection has been increasingly reported. This has been attributed to a viral relapse in a host that failed to completely eradicate the virus or to reinfection with a different viral genome (2, 3). The involvement of innate/Type I/III interferon (IFN) response as well as the specific antibody and T cell responses is well established in the protection against SARS-CoV-2 (4). However, the immune mechanisms underlying reinfection/virus relapse are still mostly unexplored. Investigation of anti-SARS-CoV-2 immunity in clinical recurrence/reinfection cases have been only directed to the humoral response. COVID-19 reinfection can occur both in the presence or absence of binding (5) or neutralizing antibodies (3, 6-9). This suggests that additional immune responses apart from neutralizing antibodies may be involved in control of reinfection and recurrence. Investigation of COVID-19 reinfection/recurrence can thus provide key information regarding immune protection mechanisms and guide vaccine studies (5).

We present a case of monozygotic twins who acquired mild COVID-19 early in 2020 and only one of them–who is a health care worker-presented recurrence four months after initial infection. In order to investigate the underlying immune mechanisms, we performed a comprehensive assessment of innate and adaptive immunity. In addition to binding and neutralizing antibody assays, we assessed the innate type I/III IFN response kinetics after TLR3 stimulation and the T cell response to SARS-CoV-2 synthetic peptides with the IFN-gamma ELISPOT assay. A second pair of monozygotic twins with concordant asymptomatic infection after exposure to COVID-19 symptomatic parents was also studied as a control.

## METHODS

### Ethics Statement

The study was approved by the Committee for Ethics in Research of the Institute of Biosciences at the University of São Paulo (CAAE 34786620.2.0000.5464) and all participants (or their parents) provided a written consent.

### Human Samples and processing

Participants were selected based on their clinical history of SARS-CoV-2 infection. Clinical data were collected in an interview and are detailed in the Supplementary Material. Participants were followed up for 6-10 months after COVID clinical symptoms and blood samples were collected on July 2020 and January 2021 for Twin pair 1 and 2, respectively. PBMCs were isolated by density-gradient centrifugation using Ficoll–Paque and stored in liquid nitrogen until use. Sera were processed and kept frozen at -80°C until use.

### TLR3 stimulus and type I/III IFN innate immune response

Cryopreserved cells were thawn and stimulated with 1µg/ml of Poly I:C HMW (Invivogen) for 1, 4 and 8h. Negative controls were incubated with R10 medium alone. Total RNA was extracted using the RNeasy Mini kit (Qiagen, Germany) and cDNA was prepared using the Superscript II Reverse Transcriptase (Thermofisher, USA), according to the manufacturer’s instructions. Real-time PCR was performed using the Power SYBR Green Master Mix (Thermofisher, USA) on a QuantStudio 12K flex (Applied Biosystems, USA). The cycling program was used as follows: 95°C for 15□min; 40 cycles of 95□°C for 15□and 60□°C for 1 min. Primers used are listed in the **Supplementary Table 1**.

### Antibody assays

SARS-CoV-2 IgG and IgM were initially detected by the clinical laboratory using the chemoluminescence immunoassay MAGLUMI 2019-nCoV IgM/IgG assay (Shenzhen New Industries Biomedical Engineering Co., Ltd, China). Enzyme-linked immunosorbent assay ELISA was performed using 96-well high-binding half-area polystyrene plates coated overnight at 4°C with 4 μg/mL of Spike protein, 2 μg/mL Nucleocapsid protein (NP) (Kindly provided by Dr Ricardo Gazzinelli, UFMG) or 0.8μg/mL of the RBD domain from human endemic coronaviruses HKU-1, OC43, NL63, and 229E, all expressed in HEK293T cells. Plasmids encoding endemic coronavirus RBD domains described in (10). Patients’ serum samples were incubated at 56°C for 30 min, diluted 1:100 and run in triplicates. In short, 50 µL of diluted sera were incubated at 37°C for 45 min. Peroxidase-conjugated goat anti-human IgG (BD Pharmingen,USA), anti-human IgA (KPL, USA) or anti-human IgM (Sigma, USA) Secondary antibody conjugates were diluted 1:10,000, and incubated at 37°C for 30 min. Values were determined as optical density (OD) minus blank and cutoff was determined as blank+ 3x standard deviation.

### Pseudovirus Neutralizing Antibody assays

The pseudovirus neutralization assay was performed exactly as described previously (11). HT1080 expressing ACE2 cells (HT1080/ACE2) and plasmids HIV-1NLΔEnv-NanoLuc and pSARS-CoV-2-SΔ19 were kindly provided by Dr. Paul D. Bieniasz (The Rockefeller University). Briefly, 104 HT1080/ACE2 were plated in 96-well plates and maintained at 37°C, 5% CO2 for 24hs. The pseudovirus was incubated in duplicate with serial dilutions of the samples for 1h at 37°C. After 48 hours of incubation at 5% CO2 at 37°C, wells were washed, and cells were lysed with Lysis Buffer (Promega). Luciferase substrate (Promega) was added to each well, and the plate read at a GlowMax luminometer (Promega). 50% inhibitory dilution (IC50) was calculated using Prism software (v7.0, GraphPad) after subtraction of the background RLUs in the control wells (cells only).

### Interferon gamma (IFNγ) ELISPOT assay

SARS-CoV2 specific T cell responses were assessed using human ex vivo IFNγ ELISPOT against a set of 20 CD4+ and 26 CD8+ T cell epitopes from 13 distinct SARS-CoV-2 proteins with high HLA allelic population coverage. We identified and synthesized the CD4+ T cell epitopes by scanning the whole proteome in SARS-CoV-2 reference genome (RefSeq: NC_045512.2) using the promiscuous HLA-DR binding peptide approach (12). The chosen CD8+ T cell epitopes were known to bind stably (www.immunitrack.com) or to be directly recognized (13) in the context of the 10 most frequent HLA class I alleles. The world population coverage of HLAs predicted to bind to the 20 CD4+ T cell epitopes and 26 CD8+ T cell epitopes was 99.6%, and 94%, respectively, according to the IEDB epitope database (14). Peptide sequences are listed in **Supplementary Table 2**. Cryopreserved PBMC were thawed and rested overnight in R10 medium (RPMI supplemented with 10% of fetal bovine serum, 2 mM l-glutamine, 1% v/v vitamin solution, 1 mM sodium pyruvate, 1% v/v non-essential amino acids, 50 U/mL Penicillin/Streptomycin and 5×10^5^ M of 2-mercaptoethanol; Thermofisher, USA) containing 30 U/ml of recombinant human IL-2 (Proleukin™, Boehringer Ingelheim Pharma, Germany). Cells were seeded at 10^5^ cells/well in MultiScreen MAIPS Filter Plates (Merck) using coating and secondary anti-IFNγ antibodies (BD Biosciences). Incubation was performed for 18 hours with synthetic peptides (5 ug/mL; Genscript), medium alone or phorbol 12-myristate 13-acetate (PMA) plus ionomycin (50 ng/mL and 1 ug/mL, respectively) and developed with AEC substrate. Spots were counted using an AID ELISpot Reader System (Autoimmun Diagnostika GmbH). The number of IFNγ producing cells/10^6^ PBMC for each peptide was calculated after subtracting the values of control wells (R10 medium alone) for each subject. The cutoff value (105 IFNγ producing cells/10^6^ PBMC) was established as the average + 3 standard deviations of test results of the 46 peptides on cryopreserved PBMC from 19 pre-pandemic Brazilian healthy control subjects (data not shown).

### HLA typing

Whole-exome sequencing was performed on peripheral blood DNA in Illumina NovaSeq platform at HUG-CELL facilities. Sequencing data was analyzed following bwa-mem and GATK Best Practices workflow, quality control and annotation was performed as previously described (15). HLA genes were realigned and called using HLA-mapper, which reduces mapping and calling errors (16).

## RESULTS

### Immunological assays in monozygotic twin pair 1 discordant for severe clinical recurrence of COVID-19

Demographical data and HLA, A, B, C and DR alleles of each of the twin pairs are compiled in **Table 1**. Clinical symptoms of COVID-19, laboratory results and dates of specific immunological tests for both twin pairs are summarized in timelines (**Figure 1**). The volunteers of the present study were identified as V01, V02 (siblings of twin pair 1), V03 and V04 (siblings of twin pair 2). For twin pair 1, initial COVID-19 symptoms were mild in both siblings (V01 in February and V02 in April). Both tested positive for anti-SARS-CoV-2 IgG in mid-May 2020. Four months after her first COVID-19-episode, V01 displayed a severe COVID-19 recurrence, with pulmonary alterations and sPO_2_<90% on room air, warranting intensive care unit admission for 24h. Her IgG titer on admission was 2-fold higher than in the first serology, in mid-May. Innate and SARS-CoV-2-specific antibody and cellular immune responses were further investigated in samples collected on August 2020 and January 2021 (**Figure 1**).

**Table 1.** Demographical data and HLA, A, B, C and DR alleles of twin pairs.

| GENERAL INFORMATION |  |  |  | HLA (Vector_CDS) |  |  |  |
| --- | --- | --- | --- | --- | --- | --- | --- |
| ID | Sex | Age range | First COVID Positive test | HLA-A* | HLA-B* | HLA-C* | HLA-DRB1* |
| V01 | F | 25-30 | IgG+ in May, 2020 | A*02:01:01;<br>A*32:01:01 | B*51:01:01;<br>B*08:01:01 | C*07:01:01;<br>C*14:02:01 | DRB1*03:01:01;<br>DRB1*11:03:01 |
| V02 | F |  | IgG+ in May, 2020 | A*02:01:01;<br>A*32:01:01 | B*51:01:01;<br>B*08:01:01 | C*07:01:01;<br>C*14:02:01 | DRB1*03:01:01;<br>DRB1*11:03:01 |
| V03 | M | 15-20 | IgG+ in July, 2020 | A*03:01:01;<br>A*23:01:01 | B*07:02:01;<br>B*35:03:01 | C*07:02:01;<br>C*12:03:01 | DRB1*14:54:01;<br>DRB1*09:01:02 |
| V04 | M |  | IgG+ in July, 2020 | A*03:01:01;<br>A*23:01:01 | B*07:02:01;<br>B*35:03:01 | C*07:02:01;<br>C*12:03:01 | DRB1*14:54:01;<br>DRB1*09:01:02 |

**Figure 1.**
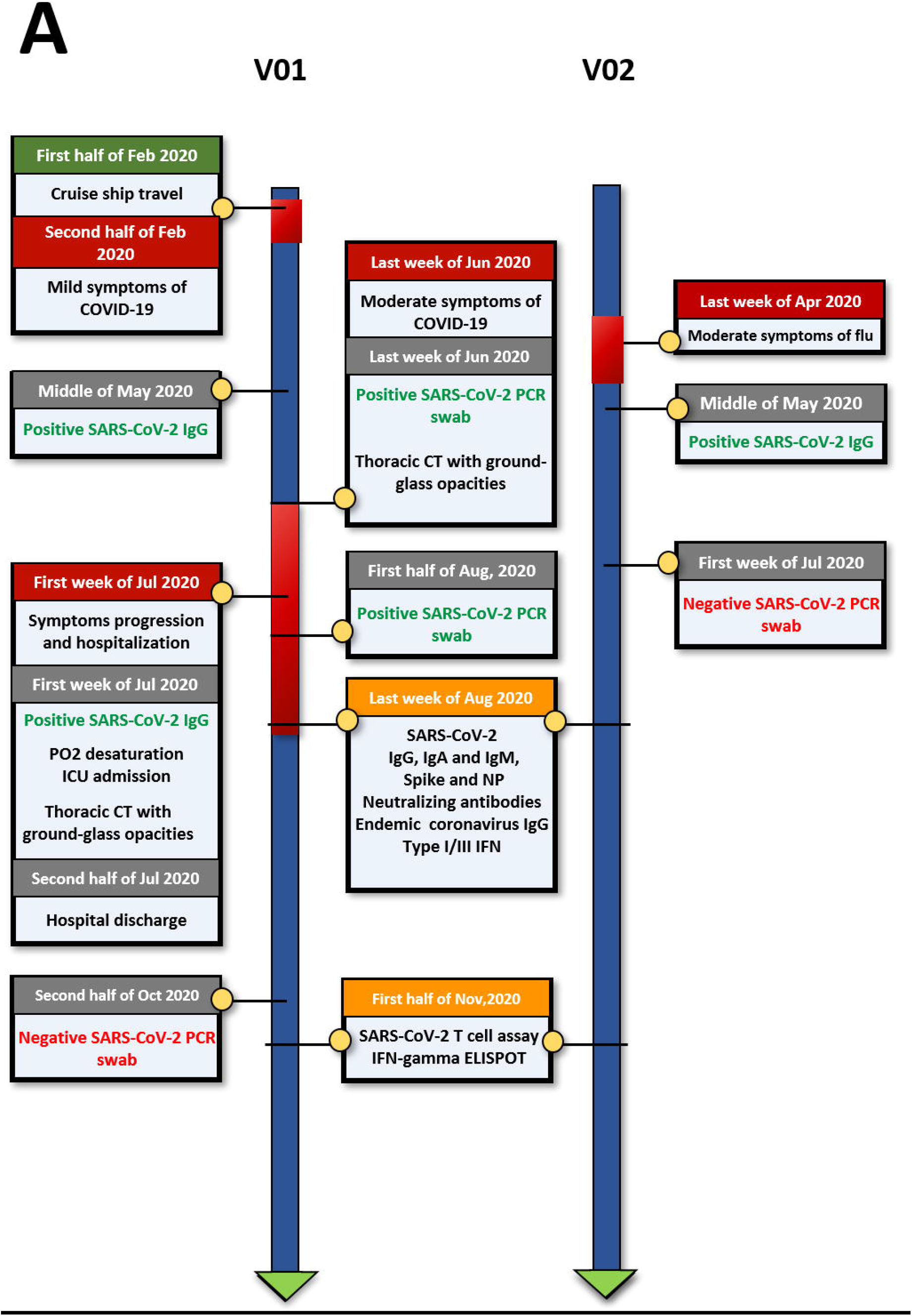
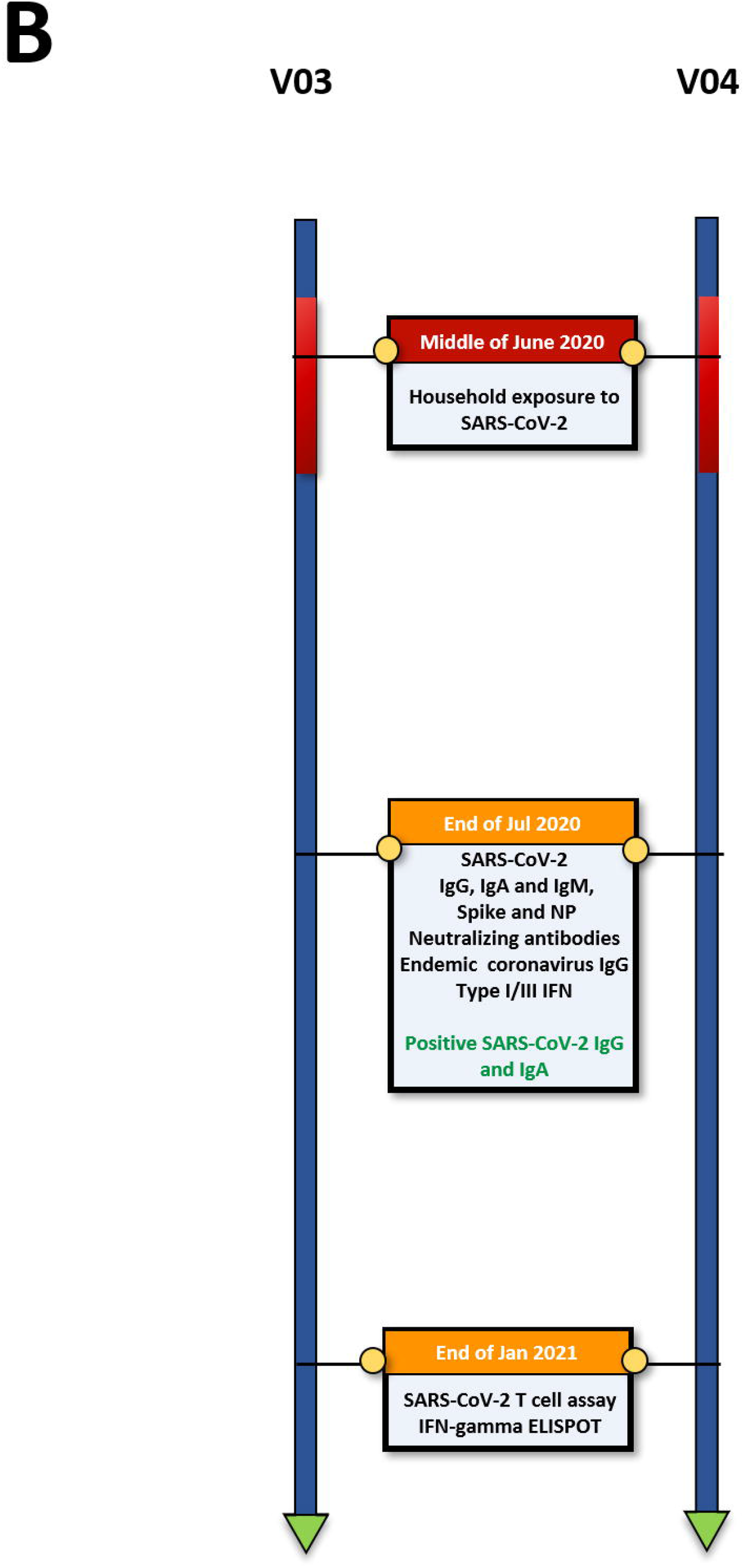
Timeline of clinical events, diagnostic results and blood draws for comprehensive immunological assessment. **A**. Pair 1 (recurrence case) and **B**. Pair 2.

Anti-Spike and anti-NP IgG, IgM and IgA antibodies were measured with ELISA (**Figure 2A**). Both twins displayed IgG against the Spike protein, with higher levels for V01 than V02 while the twin pair 2 presented anti-Spike and anti-NP IgG. Likewise, neutralizing antibodies (nAb) titers were around 1:500 for V01 and just below negative control for V02. For twin pair 2, nAb levels were similar to those of V01 (**Figure 2B**). IgG antibodies against the RBD of the four main circulating endemic coronaviruses, NL63, 229E, HKU1, OC43 were similar for all 4 cases, displaying antibodies for 3 out of 4 tested coronaviruses. The antibody profiles to human endemic coronaviruses were virtually identical in both twins V01 and V02 (**Figure 2C**).

**Figure 2.**
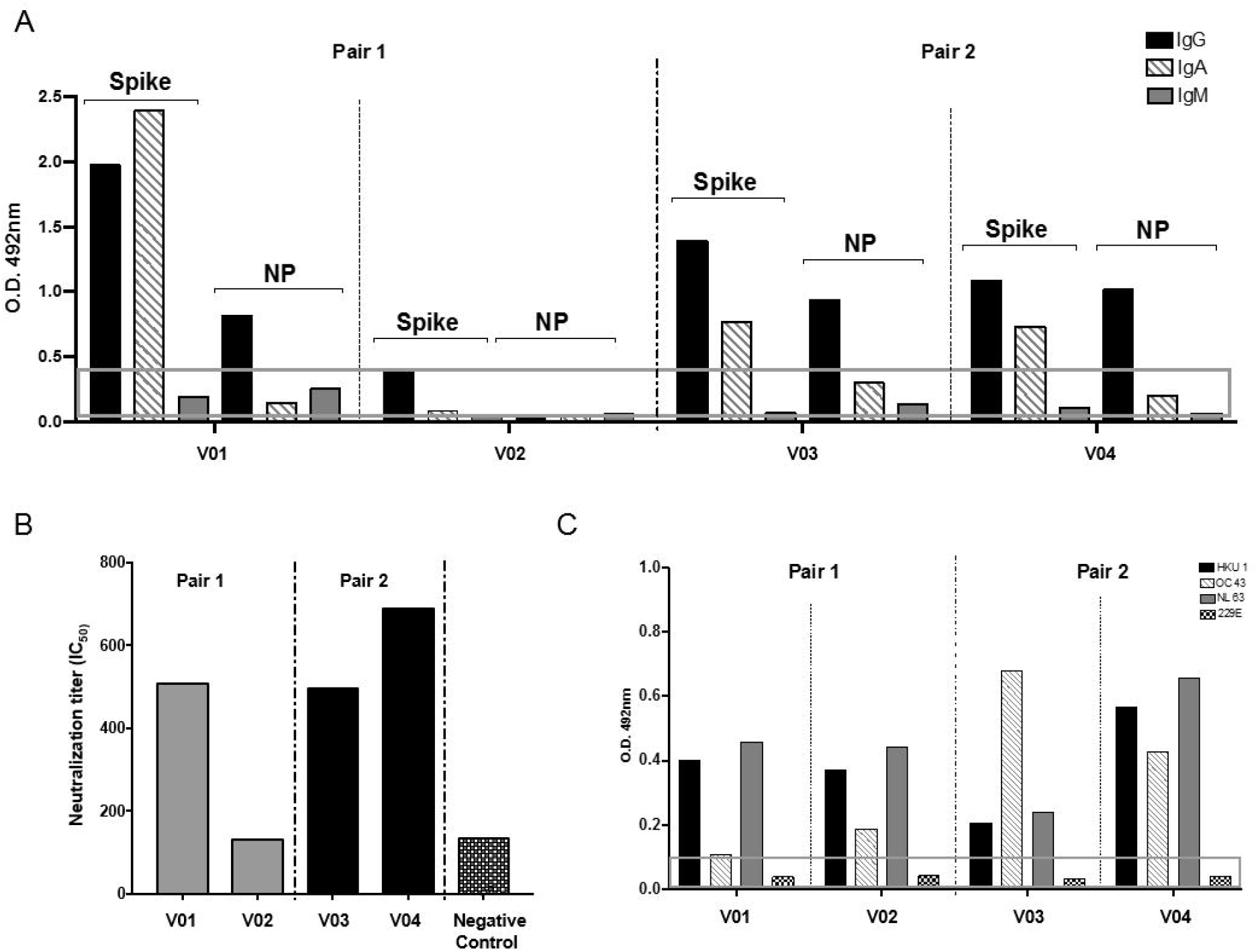
Antibody responses for SARS-CoV-2 and endemic human coronaviruses. **A**. ELISA for IgG, IgA and IgM against Spike, RBD and Nucleocapsid (NP) protein; **B**. 50% inhibitory dilution (IC50) of SARS-CoV-2 pseudovirus is showed for each individual. pair 1: V01 and V02; Twin pair 2: V03 and V04. **C**. ELISA for IgG against RBDs of human endemic alpha- and beta-coronaviruses NL63, 229E, HKU1 and OC43. The gray rectangle in **A** and **C** represent the cutoff values.

To investigate the kinetics of innate type I/III IFN responses, we stimulated PBMC with TLR stimulus (double-stranded RNA Poly I:C). We observed early (1h) and high (FC>10) mRNA expression of at least one of the four IFNs tested in all four cases. In twin siblings from pair 1, V01 expressed high IFNB1, and V02 expressed high IFNA2 and IFNL3 (**Figure 3A**). In twin pair 2, V03 expressed high IFNA2 and INFL2, while V04 expressed high IFNB1 (**Figure 3B**). Kinetics of Type I/type III IFNs was variable in later time points. Expression of ISGs was nearly identical within each twin pair, presenting an identical pattern of IFIT3, IFITM1 and IRF7 in twin pair 1 while a lower, concordant response was observed for pair 2 (**Figures 3E-H**).

**Figure 3.**
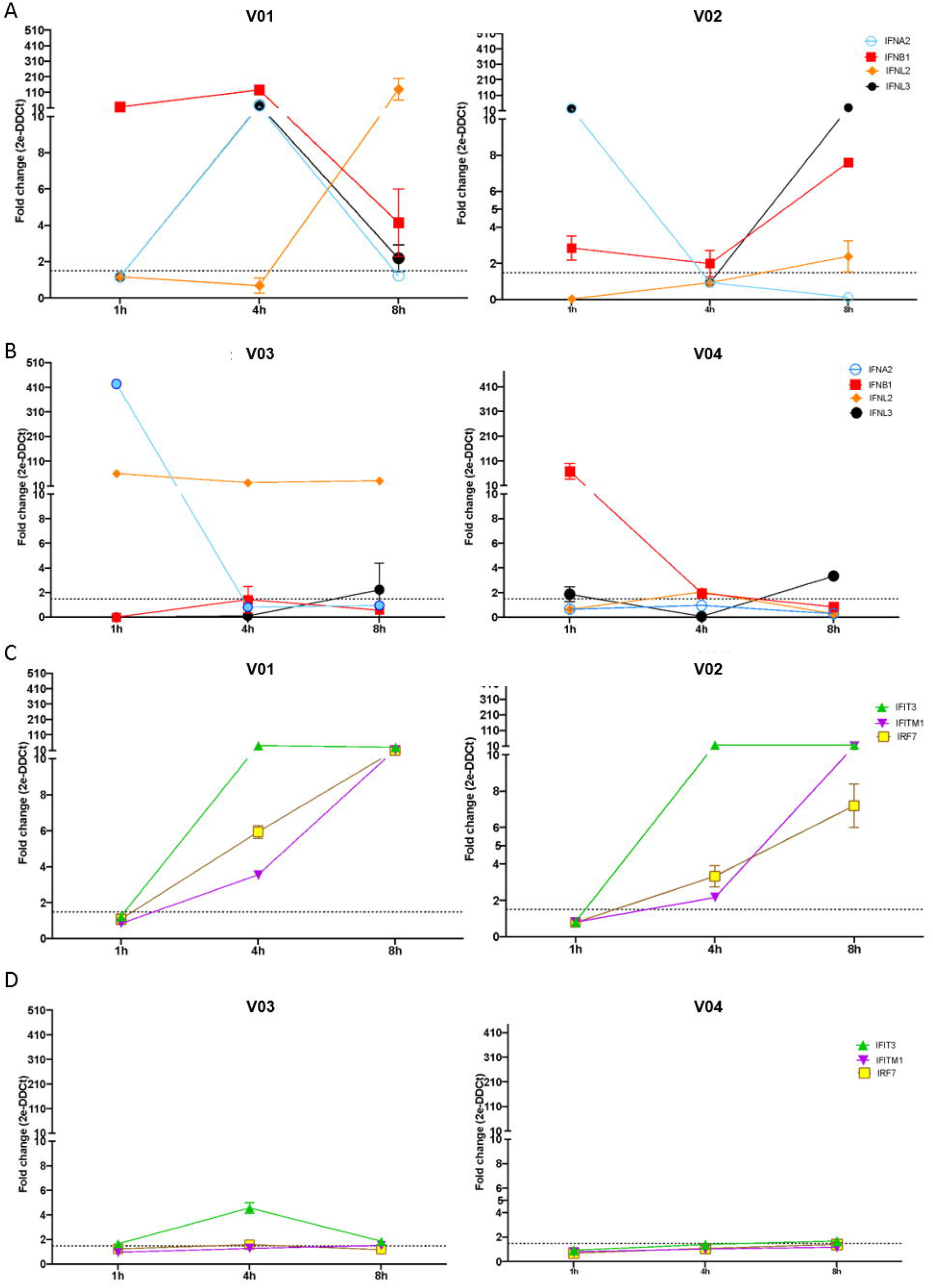
Transcriptional type I/type III IFN innate immune response. PBMCs were stimulated with 1µg/ml of double-stranded RNA Poly I:C for 1h, 4h and 8h. Total RNA was extracted for qPCR. Gene expression is relative to unstimulated cells. Expression kinetics of type I/type III genes after TLR3 stimulus for **A**. Pair 1: and **B**. Pair 2. Expression kinetics of interferon-stimulated genes (ISGs) **C**. Pair 1 and **D**. Pair 2.

To assess the T cell responses to SARS-CoV-2 T cell epitopes, we performed IFNγ ELISPOT assays on PBMC samples (see timelines in **Figure 1**). The recurrent case V01 recognized only 3 out of 20 CD4+ T cell epitopes (15%) while her sibling recognized 17 CD4+ T cell epitopes (85%; P<0.0001, Fisher Exact Test) (**Figure 4A**). Furthermore, V01 recognized only 4 CD8+ T cell epitopes (15%), while her sibling recognized 19 out of the 26 CD8+ T cell epitopes (73%; P<0.0001, Fisher Exact Test) (**Figure 4B**). Both twin siblings V03 and V04 recognized >70% of CD4+ and CD8+ epitopes (20 and 14 out of 20 CD4+ T cell epitopes and 26 and 21 out of 26 CD8+ T cell epitopes, respectively) (**Figures 4A and 4B**). These results indicate that the twin with COVID-19 recurrence showed a drastically reduced breadth (number of recognized epitopes) of both CD4+ and CD8+ SARS-CoV-2 T cell epitopes as compared with her non-recurrent sibling and also the second MZ twin pair.

**Figure 4.**
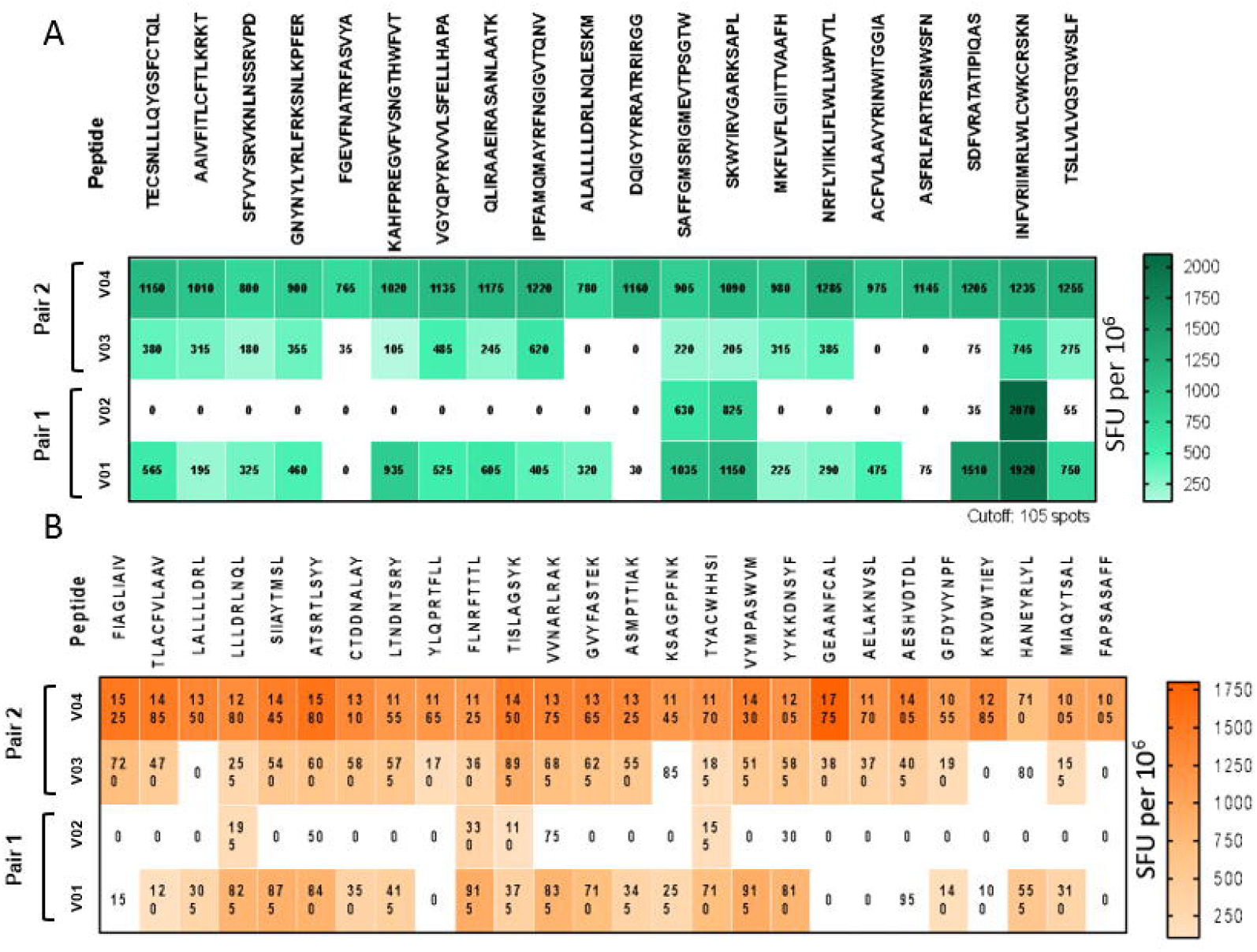
Ex vivo IFNγ ELISPOT responses to SARS-CoV-2 synthetic peptides representing **A**. CD4+ **B**. and CD8+ SARS-CoV-2 T cell epitopes. PBMC were stimulated with synthetic peptides for 18h.

## DISCUSSION

Here we describe a comprehensive assessment of innate and adaptive immune response against SARS-CoV-2 in two pairs of monozygotic twins, including a case of severe COVID-19 recurrence in one of two twin, a health care worker. We found that the only immune parameter that was substantially lower in the COVID-19 recurrence case as compared to her twin sibling and a second pair of MZ twins was the breadth (number of recognized epitopes) of the CD4+ and CD8+ T cell responses. To our knowledge, this is the first report of innate and T cell immune responses in the context of COVID-19 recurrence and reinfection. Of note, there were also fluctuations in some other immune parameters within each MZ twin pair.

Innate Type I/III IFN responses are the first line of cellular defense against RNA viruses. SARS-CoV-2 induces lower Type I/III IFN responses as compared to other respiratory viruses due to viral-encoded proteins that dampen such responses (4). An immune response characterized by a weak, delayed production of Type I/III interferons contributes to severe forms of the disease (17). The finding that all four tested patients including the COVID-19 recurrence case presented an early, strong type I/III IFN response indicates that recurrence was not associated with failure in the innate IFN response.

Protection against reinfection by viruses is mainly mediated by adaptative immune responses. Memory responses to secondary exposure to pathogen limit or prevent reinfection (5). Immunological investigation of COVID-19 recurrence and reinfection has been so far limited to antibody reactivity profiles. In a recent review of 16 reinfection cases ocurring 19-142 days after the first infection episode, 10 reported anti-SARS-CoV-2IgG testing, 6 of which displayed positive serology at the time of second infection; reinfections occurred in patients with all degrees of severity in the primary infection, but 75% of the reinfections were asymptomatic or mild (5). Neutralizing antibodies (nAb) were detected at the time of reinfection in three case reports (3, 6, 7), while only one case reported reinfection in the absence of detectable nAb (8) suggesting that responses beyond neutralizing antibodies are important to control reinfection. In our study, reinfected case V01 presented nAb 6 weeks after COVID-19 recurrence. Although we cannot ensure that neutralizing antibodies were present earlier when recurrence occurred, increased levels of anti-SARS-CoV-2 IgG were detectable at hospital admission indicating that COVID-19 recurrence boosted IgG levels, which is in line with literature reports (5).

Our finding that the CD4+ and CD8+ T cell responses of the COVID-19 recurrence case had a drastically reduced breadth 4 months after hospital discharge is indicative of a low SARS-CoV-2-specific T cell response. This is in contrast with the finding that hospitalized patients display a larger breadth of CD4+ T cell responses than outpatients (18). Given the importance of T cell responses associated with COVID-19 infection (19), a dampened CD4+ T cell response can have important consequences for many aspects of anti-SARS-COV-2 immunity. Asymptomatic and mild cases of CoVID-19 are correlated with specific CD4+ and CD8+ T cell responses, but not with IgG or neutralizing antibody, suggesting that T cells are the primary effectors controlling a primary SARS-CoV-2 infection (4, 20, 21). The dominant cytokine produced by virus-specific CD4+ T cells is IFNγ with a Th1 profile, associated with antiviral activity. CD4+ T cells protect mice from lethal SARS-CoV infection (22), and Th1 CD4+ T cells are important to provide help for the cytotoxic CD8+ T responses crucial for clearance of viral infections. CD4+ T follicular helper cells contribute for B cell responses, and IL-22-producing T cells observed in COVID-19 are key for maintenance of mucosal repair, particularly gut and lung epithelial cells (4).

It is unlikely that the reduced T cell responses observed in V01 are due the absence of HLA presentation to T cells, since her MZ twin V02 carrying the same HLA alleles displayed a broad recognition profile. Also, it is not likely that the contrasting T cell responses observed between the two siblings is a result of previous exposure to cross-reacting viruses (23), since their IgG profile against human endemic coronaviruses’ RBD was nearly identical.

Our study has limitations. We report observations for only one case of COVID-19 recurrence and cannot ensure that our findings are common in COVID-19 recurrence. We cannot conclude whether this recurrence event is a bona fide reinfection or a viral relapse in a chronic carrier since we did not genotype the SARS-CoV-2 virus. Most reinfection episodes were reported to be asymptomatic or mild (5). However, the 4-month interval between the two events in an exposed health care worker with a more severe clinical presentation is more suggestive of reinfection due to re-exposure than of a viral relapse.

In short, our results suggest that the failure in inducing a broad T cell response might have enhanced susceptibility to COVID-19 recurrence in the reported case. Our data may support a prime role for T cells in protection against reinfection. Given the increased concern that SARS-CoV-2 variants escaping antibody neutralization could give rise to a massive raise in reinfection (24, 25), our case stresses the importance of T cell immune responses in protection against reinfection. This is in line with the reported lack of deleterious effect of virus variants in the cellular immune response (26). Further investigation in a larger cohort can shed light on whether T cell dysfunction is a common mechanism for recurrence of COVID-19.

## Data Availability

Individual-level raw sequencing and immunological data can be shared upon reasonable request, as well as corresponding phenotypes.

## Author contributions

M. V. C. and K. S. S. contributed equally to this work as co-first authors. E. C. N., J.K., and M. Z. contributed equally to this work as principal investigators. All authors contributed significantly to this work.

## Acknowledgments

The authors are extremely grateful for the participation and collaboration of the four twins and their families, the nurses for samples collection, the technical team and the Fleury Laboratory for serology tests. Special thanks to JBS. S.A. for the additional funding.

